# Using digital spatial profiling to analyse onco-immune-related gene expression within oropharyngeal tumours in relation to HPV and p16 status

**DOI:** 10.1101/2023.11.17.23298621

**Authors:** Jill M. Brooks, Yuanning Zheng, Kelly Hunter, Benjamin E. Willcox, Olivier Gevaert, Hisham Mehanna

**Author notes:** Corresponding author: Hisham Mehanna, Institute of Head and Neck Studies and Education, Institute of Cancer and Genomic Sciences, University of Birmingham, Birmingham, UK. Joint senior authors (Olivier Gevaert, Hisham Mehanna). Competing interests:* KH works for Propath. HM reports advisory board fees from AstraZeneca, MSD, Merck, Nanobiotix, and Seagen and is Director of Warwickshire head neck clinic and Docpsert Health. The other authors declare no competing interests. Data availability:* Data is available on request from the corresponding author. Funding:* This work was funded by Cancer Research UK and National Institute for Health Research (NIHR) UK. The views expressed in this article are those of the authors and not necessarily those of the NIHR.

## Abstract

Human Papilloma virus (HPV)-mediated oropharyngeal cancer (OPC) has significantly better prognosis compared with HPV-negative, stimulating interest in treatment de-intensification approaches to reduce long-term sequelae. Routine clinical testing frequently utilises immunohistochemistry to detect upregulation of p16 protein as a surrogate marker of HPV-mediation. However, this does not detect discordant HPV+/p16-cases and incorrectly assigns HPV-/p16+ cases, which, given their inferior prognosis compared to HPV+/p16+, may have important clinical implications. The biology underlying poorer prognosis of HPV/p16 discordant OPC requires exploration. Here, we utilised digital spatial profiling to compare the expression patterns of selected immuno-oncology-related genes within the tumour and stromal compartments of HPV+/p16+, HPV-/p16+ and HPV-/p16-OPC tumour tissues (n=12). KRT and *HIF1A* were upregulated in HPV-/p16+ and HPV-/p16-tumours relative to HPV+/p16+. Conversely, multiple genes associated with antitumour immune responses (such as *CXCL9, CXCL10, CD74*) were upregulated in HPV+/p16+ tumours. Of note, HPV-/p16+ and HPV-/p16-tumours displayed highly similar gene expression profiles, with only CXCL9 showing differential expression in stromal regions. These results are consistent with described prognostic patterns (HPV+p16+ > HPV-/p16+ > HPV-/p16-) and underline the need for dual HPV and p16 testing to guide clinical decision making.

## Introduction

Head and neck cancer (HNC) is the seventh most common cancer worldwide^1^. Incidence rates are rising; mostly driven by a rapid increase in oropharyngeal cancer (OPC) incidence within certain global regions including the United States (US), Europe, New Zealand, and parts of Asia^2-4^. The main risk factors for OPC include smoking, excessive alcohol intake and infection with high-risk Human Papilloma virus (HPV). It is the increase in the latter which underpins rising incidence rates. In the US and United Kingdom, HPV-mediated OPC is now more prevalent than HPV-mediated cervical cancer^3,5^. It is estimated that incidence will continue to rise for the next ∼20 years before the impact of gender-neutral prophylactic vaccination is felt^6,7^.

HPV-mediated OPC has distinct epidemiological, molecular, and immunological features compared with HPV-negative disease and is associated with better treatment response and outcomes^8,9^. This has led to separate classifications in the latest UICC/AJCC TNM staging system (TNM8)^10^. Given the improved prognosis^8^ and younger age of HPV-positive OPC patients, there is considerable interest in approaches to de-intensify treatment and reduce long-term morbidity and quality-of-life impact. Unfortunately, clinical trials to date have reported limited or no success^11,12^. One potential explanation for this relates to the determination and definition of HPV status.

The presence of HPV in OPC can be assessed directly using PCR-based methods or in situ hybridisation to detect viral DNA/RNA or indirectly using immunohistochemistry (IHC) to assess overexpression of the protein p16^INK4a^ (p16)^13,14^, which is the indirect result of HPV early protein 7 (E7)-mediated inactivation of retinoblastoma protein. In contrast, frequent loss, mutation, or epigenetic silencing of the *CDKN2A* gene encoding p16 results in low/absent expression in HPV-negative OPC. Detection of p16 overexpression therefore provides a good surrogate marker for HPV status and - being simple and cost-effective to implement – is routinely used in clinical practice. However, recently dual p16 and HPV-DNA/RNA testing has shown that whilst the majority of p16-positive tumours are HPV-positive (HPV+/p16+), a subset (∼10%) are HPV-negative (HPV-/p16+). Likewise, a small subset of HPV-positive tumours do not overexpress p16 (HPV+/p16-). An important issue is whether these two discordant subsets – particularly those that are HPV-/p16+ and therefore assigned as ‘HPV-positive’ by routine testing – share the improved treatment response and survival outcomes of HPV+/p16+ cases, or whether outcomes are more closely aligned with HPV-/p16-tumours. Multiple studies suggested differential prognosis but were limited by sample size or restricted geographical sampling^15-19^. A recent large multicentre study (n = 7,654) provided strong evidence that patients with discordant OPC (HPV+/p16- or HPV-/p16+) have significantly worse prognosis compared with HPV+/p16+ OPC patients, although significantly better than HPV-/p16-patients^20^.

The biology underpinning this intermediate outcome requires exploration to develop understanding and inform clinical decision making. Here we utilised NanoString’s GeoMx Digital Spatial Profiling (DSP) platform to explore differences in gene expression between HPV+/p16+, HPV-/p16+ and HPV-/p16-oropharyngeal tumours. This approach enables spatially-resolved analysis of gene expression within defined regions of interest, selected based on expression of morphology markers; for example, pan-cytokeratin expression to identify tumour versus stroma in epithelial tumours and/or CD3 to identify T cell rich areas.

## Methods

### Cohort

The study utilised formalin-fixed, paraffin-embedded diagnostic biopsy samples from 12 OPC patients recruited to the PET-NECK^21^ or Predictr^22^ clinical studies between 2000 and 2012. Patients were treated with curative intent, with either platinum-based chemoradiotherapy or surgery followed by adjuvant radiotherapy/chemoradiotherapy. Ethical approval for use of tissue samples in translational research was granted by North West - Preston Research Ethics Committee (Reference: 16/NW/0265).

### Digital spatial profiling

5μm tissue sections were cut and mounted on SuperFrost plus microscope slides. Digital Spatial profiling (DSP) was carried out at NanoString Technologies Inc. Seattle, USA as part of their Technology Access Programme and according to their standard protocol^23^. Briefly, slides were hybridised/stained with oligo-conjugated RNA detection probes (immune-oncology panel comprising 78 genes, 73 target genes and five controls (Table 1)), plus three fluorescence-conjugated antibodies and nuclear stain for characterisation of tissue compartments to facilitate Region of Interest (ROI) selection. This utilised pan-cytokeratin (PanCK) as a tumour marker for tumour stroma segmentation, CD3 to identify T cells, CA-IX as a marker for hypoxia and Syto 83nuclear stain. Slides were imaged and a total of 141 ROIs across the 12 samples selected for analysis. ROIs were then sequentially illuminated with UV light to cleave oligo-probes which were aspirated and dispensed into 96-well plates. Probes were then hybridised to optical barcodes and counted using the nCounter platform (NanoString Technologies). Digital raw counts were exported for analysis.

**Table 1.**
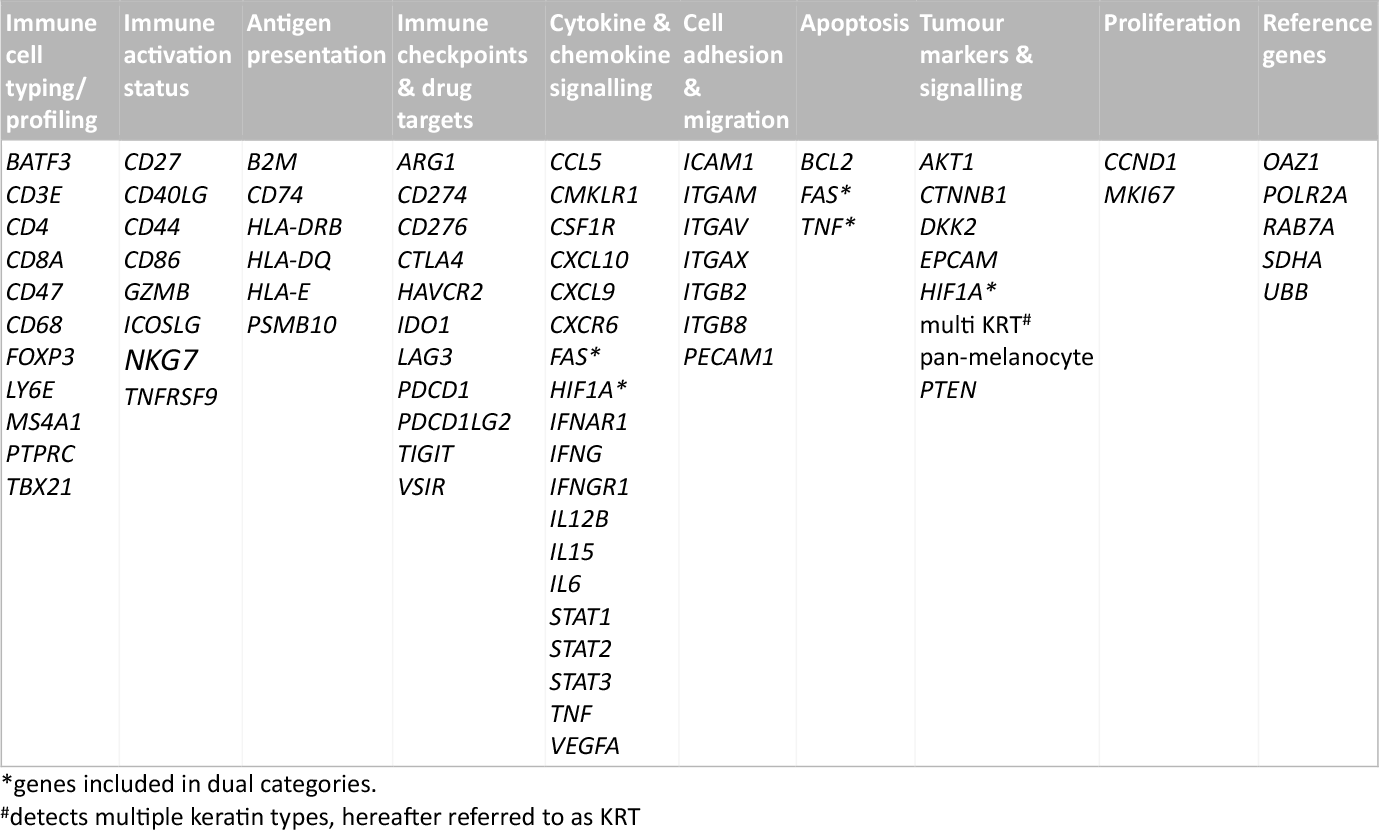
Target genes included in the GeoMx Immune-oncology human RNA panel.

### Data analysis

Quality control and normalization were performed in accordance with NanoString’s Gene Expression Data Analysis Guidelines (https://nanostring.com/wp-content/uploads/Gene_Expression_Data_Analysis_Guidelines.pdf). nCounter readout performance was assessed by evaluating the imaging and binding density QC metrics. All imaging segments demonstrated a high percentage of successfully scanned subsections, with a registered Fields of View (FOV) above 88%. The binding density was found to be less than 0.58, indicating limited overlaps between the reporter probes. Gene expression counts for the target genes were normalized to a set of house-keeping genes (RAB7A, UBB, SDHA, POLR2A, OAZ1), and a normalization factor was calculated for each segment by comparing the geometric means of house-keeping genes across all segments. The gene expression values were then log2 transformed. Differential gene expression analysis was performed using the Kruskal–Wallis test, and pairwise comparisons were made using the Mann-Whitney U test. Raw *P* values were adjusted using the Benjamini-Hochberg procedures, with adjusted *P* values below 0.05 considered statistically significant.

## Results

The cohort of 12 FFPE diagnostic biopsy samples from patients with OPC included four patients whose tumours were HPV+/p16+, three discordant (HPV-/p16+), and five HPV-/p16-. Patient characteristics are summarised in Table 2.

**Table 2:** Patient characteristics.

| Patient number | Gender | Age group | Clinical stage | HPV status<br>0 = negative<br>1 = positive | p16 status<br>0 = negative<br>1 = positive | Smoking status<br>1 = never<br>2 = past<br>3 = current | Alcohol consumption<br>1 = low<br>2 = moderate<br>3 = heavy |
| --- | --- | --- | --- | --- | --- | --- | --- |
| 2 | M | 65-70 | IV | 1 | 1 | 3 | 2 |
| 3 | M | 50-55 | NA | 1 | 1 | NA | NA |
| 4 | M | 70-75 | IV | 1 | 1 | 1 | 2 |
| 9 | M | 45-50 | IV | 1 | 1 | 3 | 2 |
| 6 | M | 55-60 | IV | 0 | 1 | 3 | 3 |
| 11 | M | 35-40 | IV | 0 | 1 | 1 | 2 |
| 12 | M | 60-65 | IV | 0 | 1 | 2 | 3 |
| 1 | M | 60-65 | IV | 0 | 0 | 3 | 2 |
| 5 | M | 65-70 | IV | 0 | 0 | 1 | 3 |
| 7 | M | 65-70 | II | 0 | 0 | 3 | 2 |
| 8 | M | 50-55 | III | 0 | 0 | 3 | 3 |
| 10 | M | 65-70 | IV | 0 | 0 | 3 | 2 |

NA – not available

Gene expression was analysed separately within the tumour (PanCK+) and stromal regions (PanCK-). ROIs were placed to capture gene expression within the tumour core, at the tumour-stroma interface and within the peri-tumoural stroma. Additional morphology markers were used to identify regions with high versus low T cell density (CDE3e) and high versus low hypoxia (as determined by CA-IX staining). Morphology marker staining, ROI placement and segmentation strategy is illustrated for one sample in **Fig. 1, A & B**. Comparison of keratin gene expression within the tumour and stromal compartments (**Fig.1C**), confirmed the efficacy of the segmentation approach.

**Figure 1.**
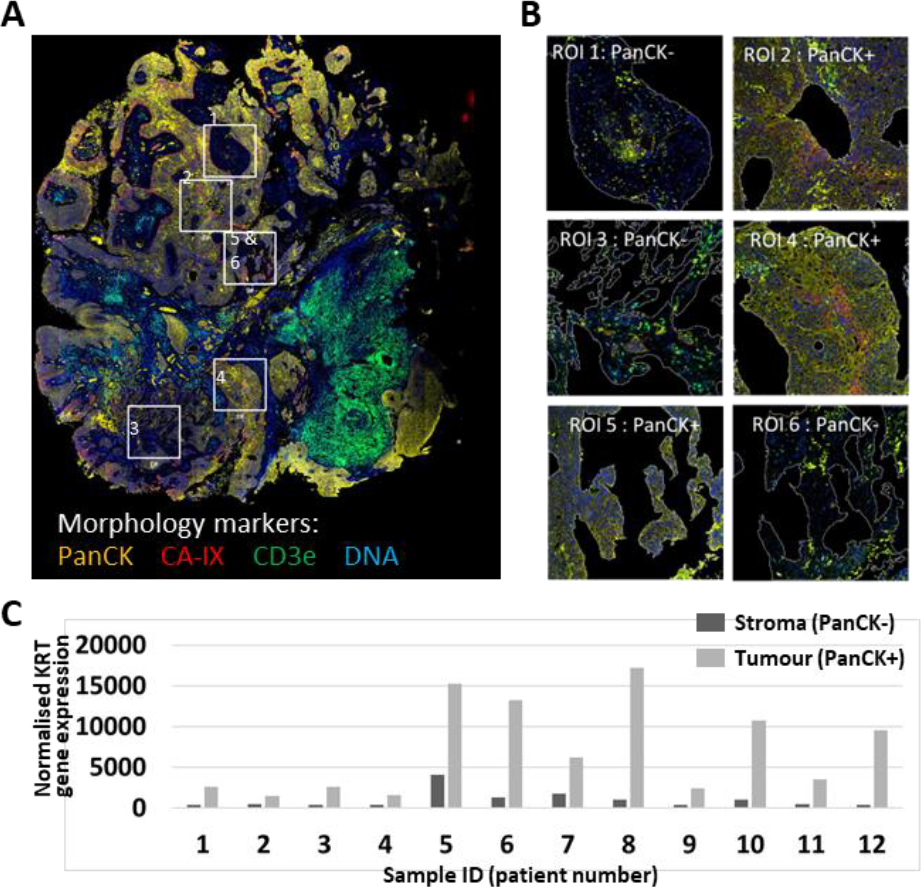
GeoMx DSP ROI selection and segmentation approach. (**A**) Tumour sections were stained with three morphology markers: PanCK (tumour; yellow), CD3e (T cell marker; green), CA-IX (surrogate marker for regions of hypoxia; red), plus a DNA stain to identify all cells. ROIs were placed to identify regions within tumour nests, tumour-stroma interface and peri-tumoural stroma. (**B**) Segmentation strategy – PanCK staining was used for identification of tumour (PanCK+) and stromal regions (PanCK-), enabling separate analysis. (**C**) Comparison of keratin gene expression between tumour and stromal compartments. Graph shows combined normalised KRT gene expression for all PanCK+ vs. PanCK-segments within each of the 12 patient samples.

Following data QC and normalisation, genes differentially expressed between the three subgroups were identified. Heat maps illustrating the top ten differentially expressed genes for each subgroup relative to the other two subgroups are shown in **Fig. 2A** and **Fig. 2B**, for the tumour and stromal compartments respectively. Pairwise comparisons of expression of all immune-oncology-related genes between the three different HPV/p16 subgroups (HPV+/p16+ versus HPV-/p16-, HPV+/p16+ versus HPV-/p16+ and HPV-p16+ versus HPV-/p16-) are illustrated in **Fig. 2C**. Key genes whose expression is down-regulated within the tumoural compartment of HPV+/p16+ versus HPV-/p16+ and/or HPV-/p16-tumours include KRT, *CD44, HIF1A, CCND1, AKT1* and *CD276* (**Fig.2C** top left and centre panels). Expression of several of these genes is also downregulated in the stromal compartment (**Fig.2C** bottom left and centre panels), as well as *ITGAV, ITGB8* and *PTEN*. Multiple genes are up-regulated within the tumour and stromal compartments of HPV+/p16+ tumours relative to HPV-/p16+ and HPV-/p16-, including several whose functions relate to antitumoral immune responses (*CXCL9, CXCL10, B2M, CCL5, CD74*, HLA-DRB, *PSMB10* and *STAT1*; **Fig. 2C** left and centre panels).

**Figure 2.**
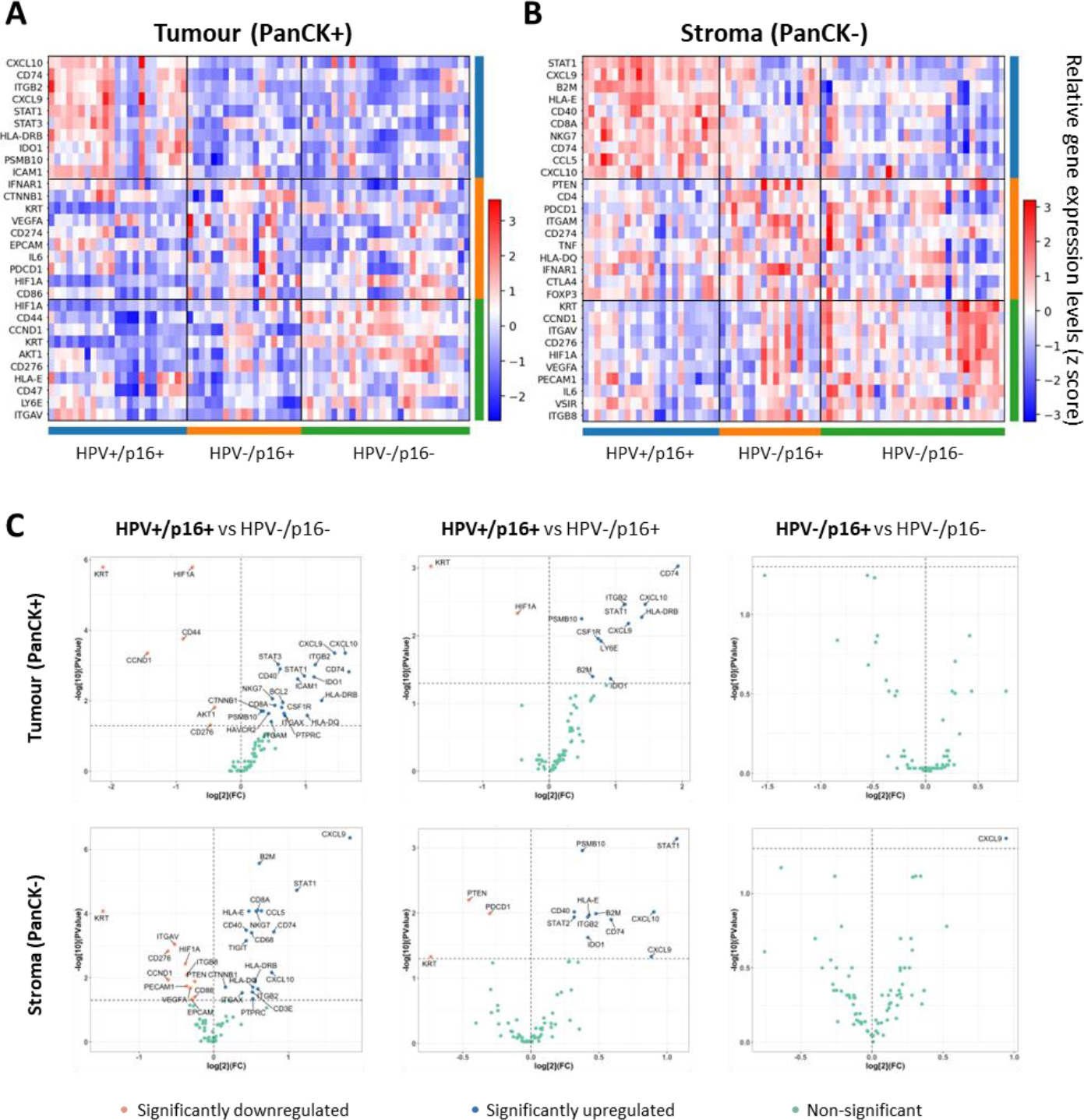
Identification of genes differentially expressed between HPV+/p16+, HPV-/p16+ and HPV-/p16-oropharyngeal tumours. (**A**) and (**B**). Heat maps showing relative gene expression levels for the top ten differentially expressed genes for each HPV/p16 subgroup within the tumour and stromal compartments respectively. Gene names are indicated for rows; each column represents an individual ROI (HPV+/p16+: n = 46; HPV-/p16+: n = 36; HPV-/p16-: n = 59. (**C**) Volcano plots showing pairwise comparisons of gene expression between the three groups for the tumour (top) and stromal (bottom) compartments. The HPV/p16 subgroup listed first is the comparator, with significantly down regulated genes shown in pink, significantly upregulated genes in blue and non-significantly altered genes in green. P values for pair-wise comparison between any two groups were calculated using the Mann-Whitney U test and adjusted by the Benjamini-Hochberg method across all genes.

Of note, in these pairwise comparisons only one gene (CXCL9) was found to be differentially expressed between HPV-/p16+ and HPV-/p16-tumours, being overexpressed in the stromal compartment of the former (**Fig.2C** right hand panels). All genes displaying significantly different expression in pairwise comparisons are listed in Supplementary Table 1.

Differential expression of selected genes (those showing the most significant differences in the pairwise comparisons) between all three groups is presented in Figure 3. The violin plots illustrate both inter-group differences in gene expression and frequency distribution of gene expression within each group. In respect of inter-group differences, Figure 3 again highlights the similarity between the HPV-/p16+ and HPV-/p16-groups and differential expression relative to the HPV+/p16+ group. With respect to frequency distribution, some genes have narrow distribution, for example, expression of KRT, *HIF1A* and *STAT1* in the stromal compartment of HPV+/p16+ tumours. Conversely, many genes have broad, bi-/multimodal distribution (for example *CD44, CCND1* and *CD74* in both tumour and stromal compartments for all three HPV/p16 subgroups), potentially reflecting spatial context, tumour core versus tumour periphery or tumour-adjacent versus more distant stroma.

**Figure 3.**
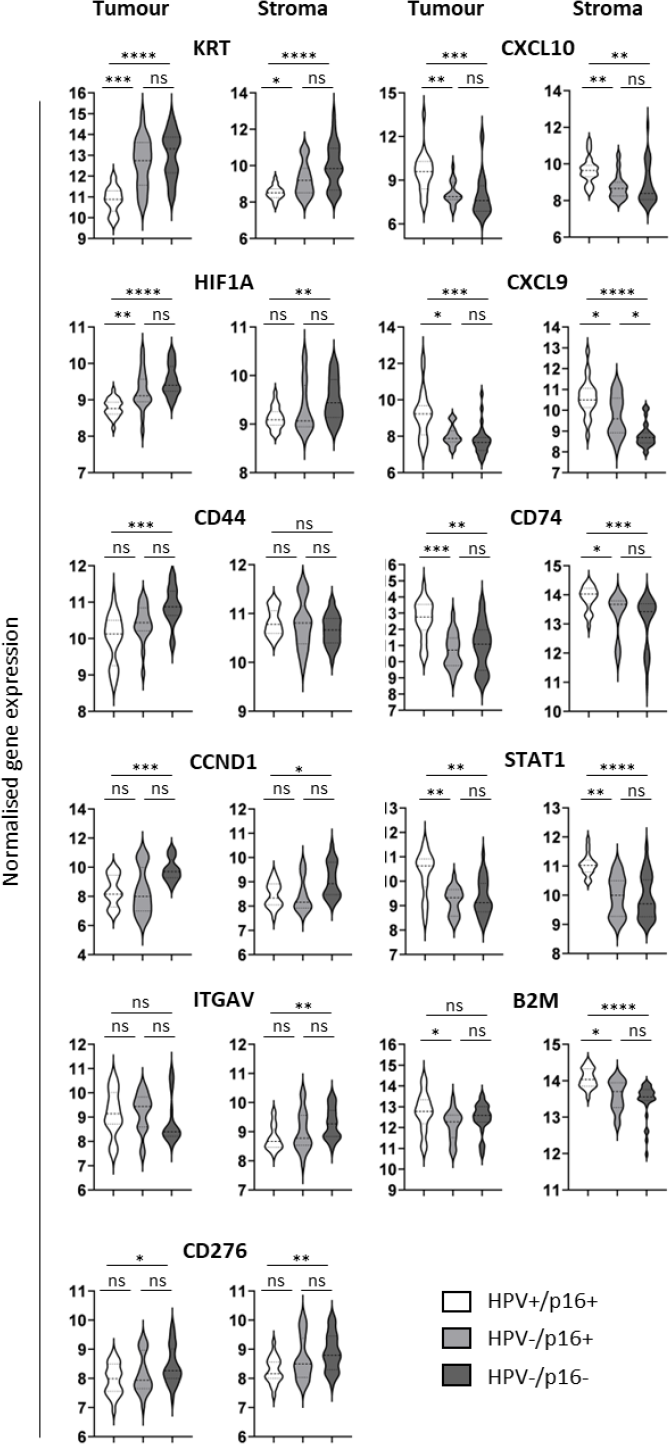
Expression of selected immune-oncology-related genes within the three HPV/p16 subgroups. Violin plots showing log2-transformed normalised gene expression for HPV+/p16+ (white), HPV-/p16+ (pale grey) and HPV-/p16-(dark grey) OPC samples. First and third columns represent tumour (PanCK+), second and fourth columns represent stroma (PanCK-). Dashed and dotted lines represent the median and quartiles respectively. Data were analysed using Kruskal–Wallis test, and pair-wise comparisons were conducted with the Mann-Whitney U test. Adjusted P values (see Supplementary Table 1) are reported as: ns, nonsignificant; *, P < 0.05; **, P < 0.005; ***, P < 0.0005; ****, P < 0.0001.

## Discussion

Spatially resolved analysis of gene (or protein) expression within distinct tumour regions offers valuable insights for unravelling tumour composition and microenvironmental phenotypes^23,24^. Here, we employed GeoMx DSP to partition OPC tissues into tumour (PanCK+) and stromal (PanCK-) compartments and quantitatively assessed gene expression within each segmented region. This approach provides valuable information that is lost in bulk RNA-sequencing. Previous studies in HNC have successfully utilised this platform to analyse the tumour and microenvironment in patients with recurrent/metastatic HNC treated with immune checkpoint inhibitors^25-27^. Here we analysed expression of immune-oncology-related genes in whole tissue sections from locally-advanced OPC patients treated with CRT +/- surgery in the curative setting, in relation to their HPV and p16 status. Overall, HPV-/p16+ and HPV-/p16-tumours showed highly similar gene expression profiles which were quite distinct from the pattern of gene expression displayed by HPV+/p16+ tumours.

The ‘gene’ showing most consistent differential expression between the three groups was KRT, being overexpressed in both HPV-/p16+ and HPV-/p16-tumours relative to HPV+/p16+ tumours. This is consistent with a previous study^28^, where overexpression of cytokeratins was observed within the HPV-negative basal HNC subtype and downregulation within the inflamed/mesenchymal HPV-positive subtype. Other studies have identified differential expression of keratin genes within HPV-positive tumours^29,30^. For example, Zhang et al identified two HPV-positive subtypes, one of which (HPV-KRT) is characterised by a complex pattern of keratin subtype up and down-regulaion^29^. In all these studies, HPV status was assigned based on detection of HPV DNA/RNA/gene signature and the effects of discordant HPV/p16 status were not explored.

*HIF1A* gene expression is upregulated in the tumoural compartment of HPV-/p16+ and HPV-/p16-tumours relative to HPV+/p16+ tumours. HIF-1α is one subunit of the heterodimeric HIF1 protein, a key transcriptional regulator of cellular adaptation to low oxygen levels. Although control of HIF-1α expression is mostly achieved through post-translational regulation (reduced degradation leading to protein accumulation under hypoxic conditions), transcriptional regulation also plays a role^31,32^. High tumoural hypoxia is associated with poor prognosis^33,34^, mediated by multiple factors including radiotherapy/chemotherapy resistance, heightened immunosuppressive nature of the tumour microenvironment, and increased epithelial-to-mesenchymal transition facilitating metastasis^35,36^. Although HPV E7 has been shown to upregulate *HIF1A* expression^37^, previous literature associates highest *HIF1A* expression with HPV-negative status^28^. Here, we identified that this association extends to include HPV-/16+ discordant tumours as well as classical HPV-/p16-, a finding that may have important implications for future treatment strategies.

Previous literature, based on phenotypic and transcriptomic analyses, describes HPV-positive tumours as ‘immune hot’^38-40^, with greater numbers of peri-tumoural immune cells and increased intra-tumoural T cell infiltration relative to HPV-negative tumours. The majority of genes upregulated in HPV+/p16+ tumours have potential roles in anti-tumoural immunity. These include key chemokines controlling T cell trafficking and infiltration (*CXCL9, CXCL10, CCL5*), components of antigen processing and presentation pathways for CD4 or CD8 T cell recognition (*B2M, PBSM10, CD74, HLA-DRB*) and STAT1, an important transcription factor promoting antitumor responses^41^. Of note, expression of these genes is similarly down-regulated in both discordant HPV-/p16+ and HPV-/ p16-tumours, excepting CXCL9 which is overexpressed in the stromal compartment of HPV-/p16+ tumours relative to HPV-/p16-. Reduced expression of genes associated with the antitumour immune response is consistent with the poorer survival outcomes of patients with HPV-/p16+ and HPV-/p16-tumours.

Our study has several limitations. Firstly, the sample size is small, reflecting the capacity of the GeoMx DSP platform which provides in depth analysis rather than high throughput screening. Secondly, the cohort does not include any examples of the second discordant group (HPV+/p16-), although this is a much smaller subgroup clinically. Thirdly, we here present analysis of 73 immune-oncology-related genes; GeoMx DSP technology has now progressed to enable whole transcriptome analysis, which we are currently employing in an extended cohort to provide even more information on mechanistic differences. For example, this might define an intermediate microenvironmental HPV-/p16+ discordant phenotype, as hinted by differential CXCL9 expression, that aligns with the observed intermediate clinical outcome.

Despite these limitations, our study clearly demonstrates that, in terms of immune-oncology-related gene expression, HPV-/p16+ OPC are more closely aligned to HPV-/p16-than HPV+/p16+, confirming the findings of a large pivotal study and underlining the need for dual testing to support informed clinical decision making.

## Data Availability

Data is available on request from the corresponding author.

---

**Supplementary Table 1.**
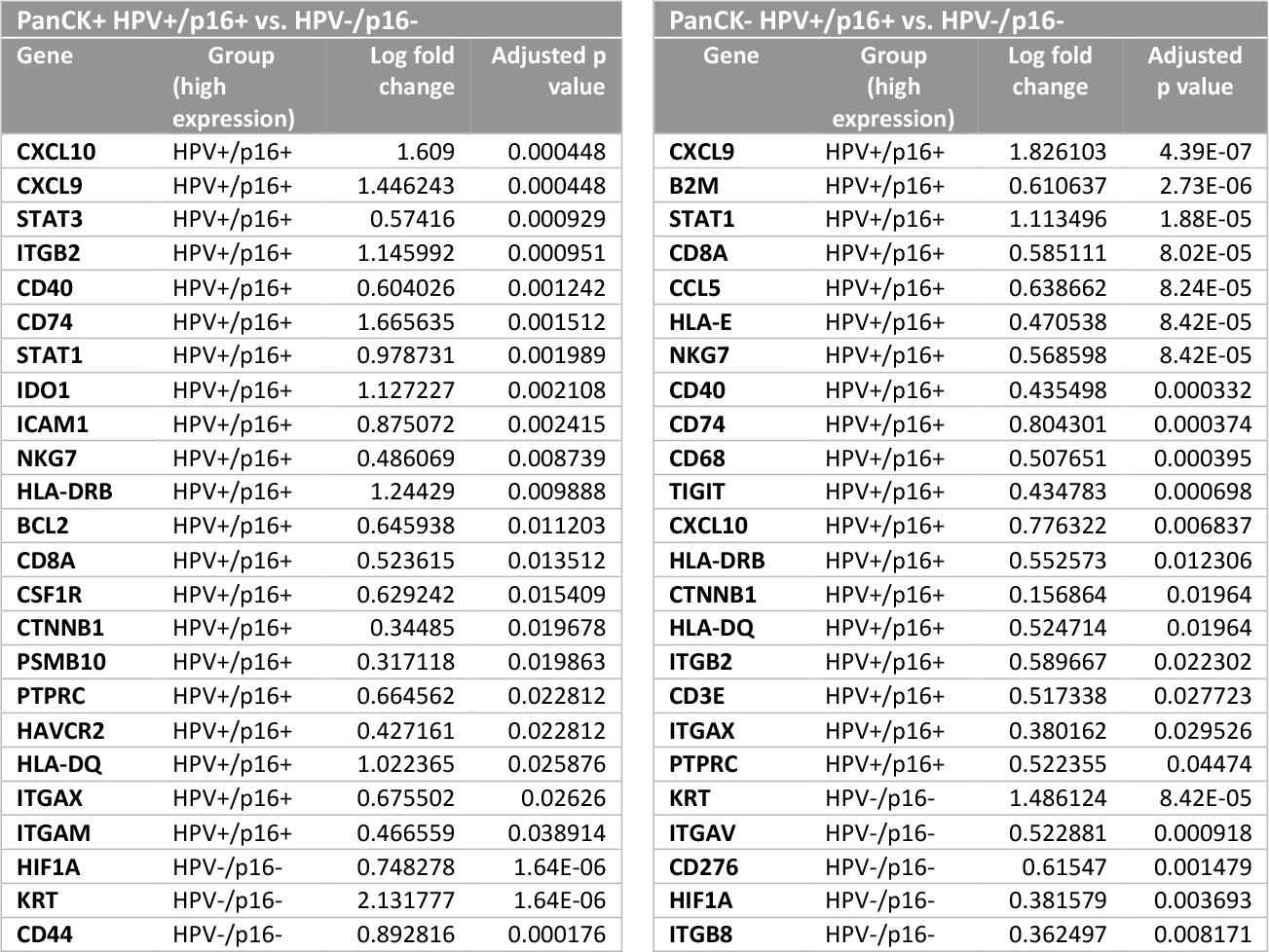
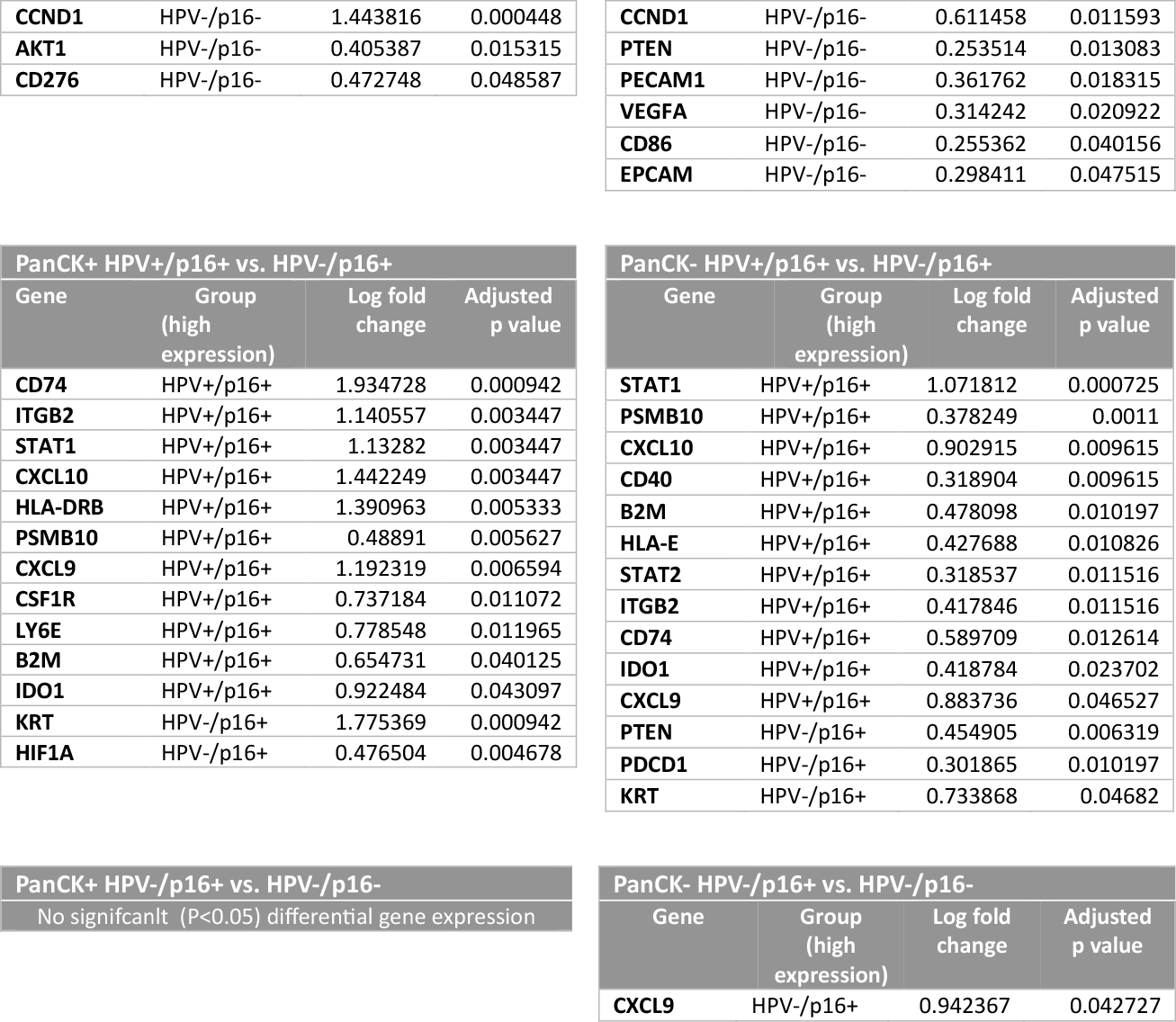
Genes displaying significantly different expression in pairwise comparisons between the three groups. Left hand columns: tumour (PanCK+); right hand columns stroma (PanCK-)

## Notes

### Author Declarations

Ethical approval for use of tissue samples in translational research was granted by North West - Preston Research Ethics Committee (Reference: 16/NW/0265).

